# Dual Attention Multiple Instance Learning with Unsupervised Complementary Loss for COVID-19 Screening

**DOI:** 10.1101/2020.09.14.20194654

**Authors:** Philip Chikontwe, Miguel Luna, Myeongkyun Kang, Kyung Soo Hong, June Hong Ahn, Sang Hyun Park

## Abstract

Chest computed tomography (CT) based analysis and diagnosis of the Coronavirus Disease 2019 (COVID-19) plays a key role in combating the outbreak of the pandemic that has rapidly spread worldwide. To date, the disease has infected more than 18 million people with over 690k deaths reported. Reverse transcription polymerase chain reaction (RTPCR) is the current gold standard for clinical diagnosis but may produce false positives; thus, chest CT based diagnosis is considered more viable. However, accurate screening is challenging due to difficulty in annotation efforts of infected areas, curation of large datasets, and the slight discrepancies between COVID-19 and other viral pneumonia. In this study, we propose an attention-based end-to-end weakly supervised framework for the rapid diagnosis of COVID-19 and bacterial pneumonia based on multiple instance learning (MIL). We further incorporate unsupervised contrastive learning for improved accuracy with attention applied both in spatial and latent contexts, herein we propose Dual Attention Contrastive based MIL (D*A*-CMIL). D*A*-CMIL takes as input a several patient CT slices (considered as a bag of instances) and outputs a single label. Attention based pooling is applied to implicitly select key slices in latent space, and spatial attention learns slice spatial context for interpretable diagnosis. A contrastive loss is applied at the instance level to encode similarity in features from the same patient against pooled patient features. Empirical results show our algorithm achieves an overall accuracy of 98.6% and an AUC of 98.4%. Moreover, ablation studies show the benefit of contrastive learning with MIL.

## 1. Introduction

The coronavirus disease 2019 (COVID-19), first recognized in Wuhan, China has spread to a global scale infecting millions and causing death to hundreds of thousands. As of August, 2020 infections surpassed 18 million, with reported deaths reaching over 690,000 globally. Caused by severe acute respiratory syndrome coronavirus 2 (SARS-CoV-2), COVID-19 is highly contagious with increasing infections each day. Despite having a relatively lower fatality rate Mahase (2020) than SARS and Middle East Respiratory Syndrome (MERS), COVID-19 has already caused more deaths. Consequently, there is an urgent need for rapid diagnosis to improve prevention while an effective vaccine is being developed.

Reverse transcription polymerase chain reaction (RTPCR) is the current gold standard for COVID-19 diagnosis based on viral nucleic acid (VNA) Zu et al. (2020). However, low sensitivity, high number of false positives and lengthy test to diagnosis times pose a challenge for early identification and treatment of patients Ai et al. (2020). Moreover, potential patients left unattended increase the risk of spreading the infection. As an easy non invasive imaging alternative, chest computed tomography (CT) is viable for fast diagnosis Ai et al. (2020). It can detect key imaging characteristics manifested in infected areas such as ground glass opacity (GGO), multifocal patchy consolidation and/or bilateral patchy shadows Wang et al. (2020a). However, image characteristics between COVID-19 and other pneumonia types may possess similarities, making accurate diagnosis challenging. Also, automated screening sensitivity is limited and not on par with radiologist level performance Wang et al. (2020b). Therefore, there is an urgent need to improve and/or develop robust screening methods based on chest CT.

On the other hand, deep learning LeCun et al. (2015) based solutions have shown success in medical image analysis Litjens et al. (2017) due to the ability to extract rich features from clinical datasets, and include a wide range of application areas such as organ segmentation Ronneberger et al. (2015) and disease diagnosis, etc. Deep learning has been employed for the diagnosis of COVID-19 in chest CT Song et al. (2020); Gozes et al. (2020a,b) and community acquired pneumonia (CAP) Kermany et al. (2018). For example, Ouyang et al. (2020) proposed a 3D convolutional neural network (CNN) with online attention refinement to diagnose COVID-19 from CAP and introduced a sampling strategy to mitigate the imbalanced distribution of infected regions between COVID-19 and CAP. Song et al. (2020) proposed DeepPneumonia for localization and detection of COVID-19 pneumonia; attention was also applied to detect key regions with impressive results on a large cohort. Despite showing promising performance, most methods are supervised and require considerable labeling efforts. Notably, even without annotated examples of infection areas, some works use existing deep learning models Shan et al. (2020) to extract infection regions and/or manually select slices in CT that show key characteristics. However, taking into consideration that during the pandemic experts have had limited time to perform labeling of CT volumes for supervised methods, unsupervised or weakly supervised learning methods that do not heavily rely on extensive data pre-processing and/or strong prior knowledge are a preferred option for accurate diagnosis.

Recently, several works focused on accurate diagnosis under weak supervision have been proposed. Notably, we consider approaches that use (a) patch-based Jin et al. (2020); Shi et al. (2020b) (b) slice-based Gozes et al. (2020b,a); Hu et al. (2020), and (c) 3D CT-based Han et al. (2020); Zheng et al. (2020) methods for diagnostic decisions. The first often uses prior segmented infection regions as input to train classifiers in a two-stage setup. The second performs slice-wise inference directly, whereas 3D based methods use the entire 3D CT scans as input with 3D convolutional neural networks (CNN). For the patch and slice-based approaches to be effective, infected regions must be well selected for training. Also, 3D CNN models are inherently slow during inference due to bigger model size and may lack interpretability.

In this work, we propose a novel end-to-end attention-based weakly supervised framework using multiple instance learning (MIL) Carbonneau et al. (2018) and self-supervised contrastive learning Chen et al. (2020a) of features towards accurate diagnosis of COVID-19 from bacterial pneumonia. We refer to this framework as D*A*CMIL. The goal of D*A*-CMIL is to assign patients a single category label i.e. (COVID-19 or bacterial pneunomia) given as input a CT volume of multiple 2D slices. In general, each patient CT scan is considered as a bag of instances that may be positive or negative. Moreover, it would be beneficial to identify which slices/instances contribute to the final patient diagnosis with the potential to localize infected regions. Herein, we propose a permutation-invariant attention based MIL pooling of slices to obtain a single representative feature for patients. In addition, spatial attention is jointly applied to learn spatial features key for infection area discovery. We incorporate contrastive learning at the instance level to encourage instance features from the same patient to be semantically similar to the patient level aggregated features in an unsupervised manner. To achieve this, an unsupervised contrastive loss is employed alongside patient category labels for the supervised loss during training.

Existing works using MIL applied in different domains often decouple instance and bag level learning into a two-step procedure i.e. first learn instance level encoders, then learn aggregation models for inference using trained encoders with MIL pooling Hashimoto et al. (2020); Hou et al. (2016). However, due to ambiguity of the instance labels and noise, learning a robust encoder can be challenging. Herein, the proposed framework aims to address the aforementioned challenges via end-to-end learning; instance selection is implicity achieved via attention based pooling of CT slices with model optimization focused only on accurate patient labels. Moreover, by jointly using supervised and constrastive loss, our model can avoid overfitting when trained on smaller datasets and improve feature robustness at the instance level without sacrificing accuracy. We empirically show the benefit of D*A*-CMIL on a recently collected dataset, with interpretable results and competitive performance against state-of-the-art methods.

The main contributions of this study include:

- We propose a novel end-to-end model for weakly supervised classification of COVID-19 from bacterial pneumonia.
- We show that joint contrastive learning of instance features and patient level features in the MIL setting is viable. A novel setting of learning instance level features without inferring labels.
- Towards interpretability, we show that dual attention, in particular spatial attention can be used to assess and visualize model decisions.
- We empirically show that D*A*-CMIL is robust to different CT sizes when instance (i.e. slice/patch) count varies via ablation studies.

The rest of the article is arranged as follows. In Section 2, we review recent works related to computer aided diagnosis with artificial intelligence for COVID-19 and relevant methodologies under weak supervision. We introduce the relavant background and details regarding D*A*CMIL in Section 3. In Section 4, we provide descriptions on experimental settings and datasets employed. Experimental results are discussed in Sections 5 & 6. We conclude this study in Section 7.

## 2. Related Works

This section presents related works in terms of COVID-19 screening, methods for weak supervision and self-supervised learning.

### 2.1. Deep Learning for COVID-19 diagnosis

The success of deep learning based techniques applied to medical image analysis has shown promising results for several application areas such as segmentation and disease detection. Several pioneering methods Shi et al. (2020a); Gozes et al. (2020a); Xie et al. (2020); Jin et al. (2020) have been proposed for the analysis of COVID-19 in both X-ray and CT images. COVID-19 lesion segmentation Gozes et al. (2020a); Xie et al. (2020), automated screening Jin et al. (2020); Song et al. (2020); Han et al. (2020) and severity assessment Huang et al. (2020) have been key areas of research. Notably, a recent review Shi et al. (2020a) shows that screening is predominant and continues to receive much interest. Moreover, given that chest CT best shows key image characteristics for COVID-19 diagnosis, CT is preferred over X-ray despite it being a low cost solution. Ng et al. (2020) recently claimed that consolidative and/or ground glass opacities (GGO) on CT are often undetectable in chest radiography and highlighted the pros and cons of each imaging modality.

Accordingly, Oh et al. (2020) recently proposed a patch-based CNN for COVID-19 diagnosis applied to chest radiography with limited datasets. They show that statistically significant differences in patch-wise intensity distributions can serve as biomarkers for diagnosis of COVID-19; with existing correlations to current radiological findings of chest X-ray. Alom et al. (2020) introduced a multi-task deep model that jointly considers chest CT and X-ray for diagnosis. They showed impressive results in both modalities for both detection and localization of infected regions. Song et al. (2020) developed DeepPneumonia, a deep learning system with rapid diagnosis to aide clinicians. Mei et al. (2020) used deep learning to integrate chest CT findings with clinical information such as laboratory tests and exposure history to rapidly diagnose COVID-19 patients. From a technical standpoint, most methods require pre-segmented lesions prior to training, and/or include multi-stages in the frameworks. Moreover, patch-based methods may suffer from noisy samples in scans, often requiring careful manual selection of slices for efficiency.

### 2.2. Weak Supervision and Multiple Instance Learning

MIL is a form of weakly supervised learning where labels/categories are provided only for a bag of instances i.e. training instances arranged in sets and the labels of instances contained in the bags are unknown Carbonneau et al. (2018). In this study, we consider a patient CT scan as a bag with unlabeled slices (instances), having only the diagnostic label for training. In general, existing algorithms can be categorized as instance-level Hou et al. (2016), bag-level Hashimoto et al. (2020), embedding-based, and joint methods that combine several approaches such as attention mechanisms Hashimoto et al. (2020); Ilse et al. (2018); Han et al. (2020). In literature, MIL has been applied to several domains including object detection Zhang et al. (2016a), image classification Yao et al. (2019); Hou et al. (2016); Zhang et al. (2016a), and object tracking Hu et al. (2017).

Also, several works have been applied in the medical imaging domain Zheng et al. (2020); Hu et al. (2020); Han et al. (2020); Wang et al. (2020c); Campanella et al. (2019). Hashimoto et al. (2020) recently introduced a novel CNN for the classification of malignant lymphoma in histopathology slides. Notably, they combined domain adaptation and multi-scale approaches with MIL for improved performance. Ilse et al. (2018) proposed attention-based pooling for MIL in an end-to-end framework; impressive results are shown across different domain problems including cancer region detection in histopathology. Weakly supervised detection of COVID-19 infection regions in chest CT is presented by Hu et al. (2020) with multi-scale learning applied for localization. More recently, Wang et al. (2020c) proposed (DeCoVNet), a method applied to 3D CT volumes using 3D CNNs with weak labels. DeCoVNet takes as input a CT volume and its lung mask for COVID-19 classification. Han et al. (2020) proposed AD3DMIL, a 3D MIL method with attention for COVID-19 screening with a deep instance generation module based on 3D latent features inspired by the pioneering work of Feng and Zhou (2017).

### 2.3. Self Supervised Learning

Self Supervised Learning (SSL) is a form of unsupervised learning where the data provides the supervision, and the network is trained to solve auxiliary tasks with a proxy loss. This is highly beneficial, especially in medical imaging where supervision is limited and the existing difficulty of curating annotations. Auxiliary tasks include context prediction Oord et al. (2018), automatic colorization Zhang et al. (2016b), and image inpainting Pathak et al. (2016). Most recently, Chen et al. (2020a) introduced a simple framework for contrastive learning (Sim-CLR) that uses extensive data-augmentation for defining predictive tasks, which achieves comparable performance to state-of-the-art supervised methods. For medical imaging tasks, He et al. (2020) recently proposed (Self-Trans) a method that combines contrastive self-training with transfer learning for COVID-19 diagnosis. Notably, the authors focus on establishing robust strategies for transfer learning with limited data, and/or when using external datasets for COVID-19 Chest CT analysis.

Inspired by recent works both for MIL and SSL, we propose to synergistically integrate contrastive self-supervision with MIL in an end-to-end framework. Notably, though previous works such as AD3DMIL Han et al. (2020) have shown impressive results; the model is based on 3D CNN and considerably has a larger model size. Also, Self-Trans He et al. (2020) follows a two-step approach by first pre-training the network via self-supervision using Chen et al. (2020b); then performs fine-tuning or transfer learning, joint self supervised training with transfer learning is underexplored. Thus, we aim to extend the current scope of the literature regarding COVID-19 via a novel formulation of MIL and self-supervised contrastive learning.

## 3. Methods

This sections presents the necessary notations and overall objectives of the task of COVID-19 diagnosis, including details of the relative modules of the proposed method.

### 3.1 Preliminaries

We consider a chest CT dataset 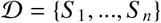 where the model receives a set of *m* labeled example scans 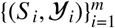 drawn from the joint distribution defined by 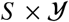. *S_i_* is a patient CT scan with instances (i.e. 2D CT slices or patches) and 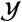 is the label set of patient-level labels, wherein 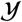 is {0, 1} for binary classification of COVID-19 and other. Also, *S_i_* is considered as a bag of instances with *S_i_* = {*s*_1_, *s*_2_,…, *s_N_*} where *N* denotes the total number of instances in the bag. It can be assumed that each instance *s_n_* has a label *y_n_* ∈ {0, 1}, however not all instances may be negative or positive. Moreover, not all slices in a scan may show infection regions vital for diagnosis, as others may be noisy artifacts not useful for learning.

Accordingly, MIL must satisfy the following constraints: if a bag *S_i_* is negative, then it can be assumed that all corresponding instances should be negative. In the case of positive bags, at least one instance is assumed to be positive. Formally, it follows that

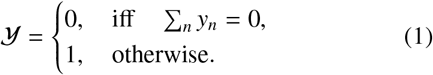

In this work, this assumption may not hold given that both sets of bags (COVID-19 and other pneumonia) considered contain both negative and positive instances (lesions). Thus, we consider a relaxed version of this constraint wherein an attention mechanism is applied to implicitly weight instances and learn their labels.

### 3.2 Proposed Approach

We developed a CNN model for patient CT scan level diagnosis between COVID-19 and other pneumonia in a single end-to-end framework. Herein, a dual-attention multi-instance learning deep model with unsupervised contrastive learning (D*A*-CMIL) is proposed. As presented in Figure 1, our method takes a CT scan with unlabeled instances as input and learns key semantic representations. It further uses an attention-based pooling to transform patient instances into a single bag representation for final prediction (see Section 3.3). Unsupervised contrastive learning is employed to encourage instances in a bag to be semantically similar to the bag representation during training (see Section 3.4).

**Figure 1:**
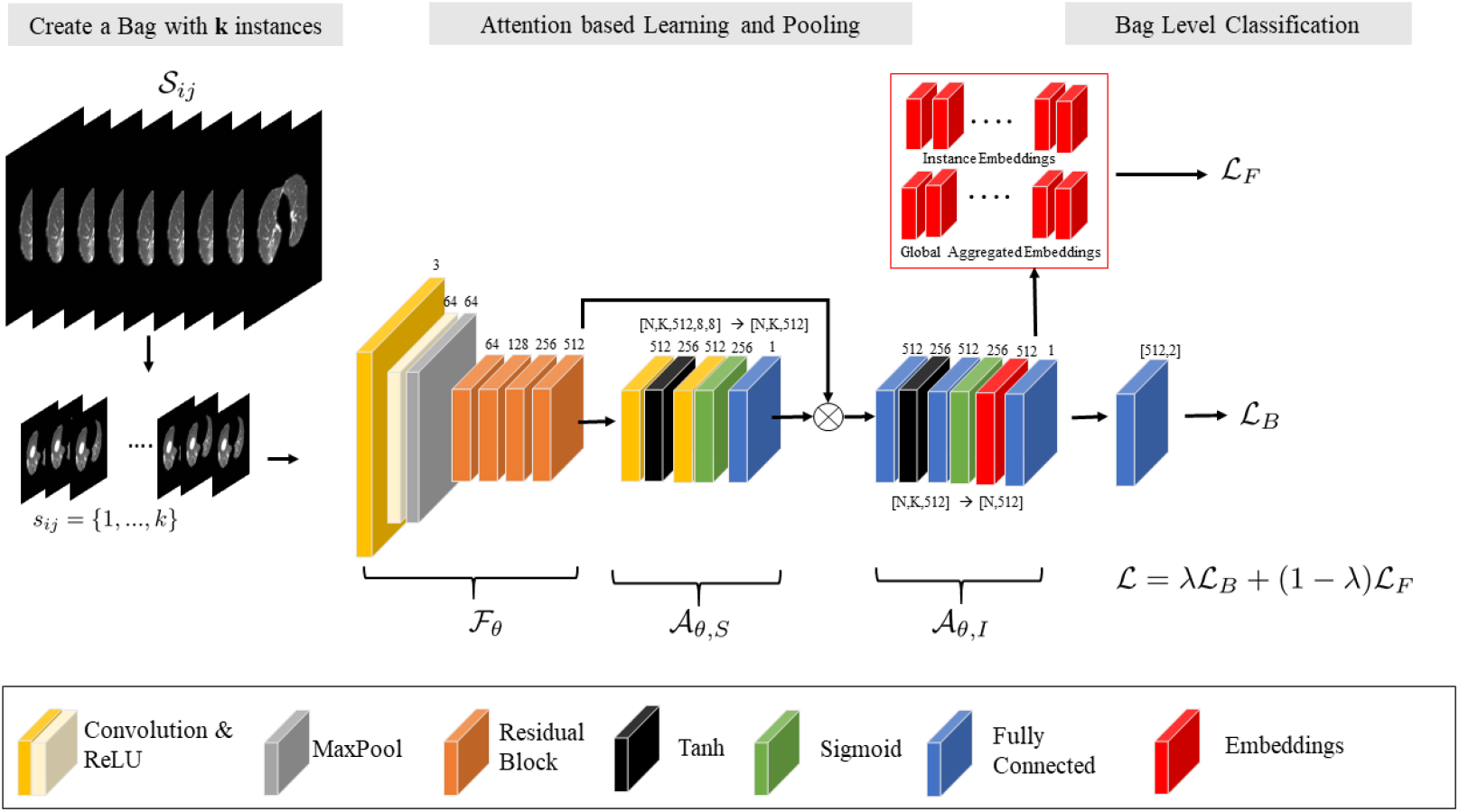
Overview of the proposed framework. For a given patient CT scan, we sample *k* instances during training to create a bag as input and feed them through the backbone *F*_θ_ to obtain feature maps. Modules *A*_θ,*S*_ and *A*_θ,*I*_ learn spatial and instance attention, then perform permutation invariant pooling via *A*_θ,*I*_ on feature maps to obtain a single bag representation. Prior to pooling, attention-weighted instance features from *A*_θ,*I*_ are employed for unsupervised contrastive learning as well as patient level learning to obtain final predictions and update the model.

In the proposed framework, a backbone CNN model 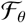 is implemented as a feature extractor to transform *i*-th instance from a CT bag into a low dimension embedding 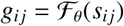 with spatial dimensions of shape *C* × *H* × *W*, where *C, H* and *W* are channel size, height and width, respectively. Then, a spatial attention module *A*_θ,*S*_ is employed to learn spatial representative features in the instances and output spatial attention maps 1 × *H*^*^× *W*^*^with *C* = 1; used to weight all instances to obtain a single spatial pooled feature 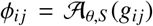, with 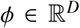, where *D* is the feature dimension size. To aggregate the instance features φ_*n*_ for each CT scan, we implement a second module 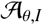 that performs attention-based permutation invariant pooling to obtain a single bag representation 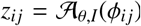, with 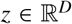 having the same dimension for consistency. Following, *z_n_* is passed to the patient level classifier 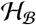 to obtain predictions for the entire bag 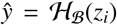, where *ŷ* is the probability of a CT scan being labeled as COVID-19 or other pneumonia. Formally, we employ the bag loss 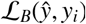 using cross-entropy. It follows that

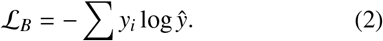

### 3.3 Dual Attention based Learning

In recent works Hashimoto et al. (2020); Ilse et al. (2018); Han et al. (2020) attention has shown to be vital for learning robust features, especially under the MIL setting. In particular, attention-based poolingIlse et al. (2018) is preferred over existing pooling methods such as max or mean, since they are not differentiable/applicable for end-to-end model updates. In this work, we implement both spatial (*A*_θ,*S*_) and latent embedding (*A*_θ,*I*_) based attention pooling via the respective modules. In the spatial module, given the input 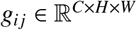, we employ two convolutional layers each followed by hyperbolic tangent (tanh) and sigmoid (sigm) non-linearities, respectively. Feature maps are passed to each module successively, then to the final convolutional layer with single channel output representing the presence of infection. Notably, we perform element-wise multiplication between the output of each branch of the convolutional layers before passing to the final layer to obtain spatial scores 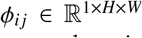. It is worth noting that we simply implement gated spatial attentionDauphin et al. (2017) instead of the commonly applied global average pooling (GAP) on the final backbone features *g_n_*.

In order to aggregate the features φ_*n*_, we employ attention based pooling proposed by Ilse *et al*Ilse et al. (2018) in the instance attention module *A*_θ,*I*_. Formally, we consider the same architecture applied for gated spatial attention, except all convolutional layers are replaced with fully connected layers since attention is applied to instance embeddings. We denote *H* = {φ_1_, φ_2_, φ_3_,…, φ_*N*_}, with *h_i_* ∈ *H^N^* as a bag with *N* instance features. Then, attention based pooling MIL with gating mechanism is defined as

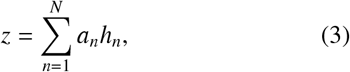

with,

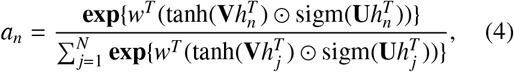

where 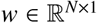, 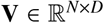, and 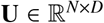 are trainable parameters. *tanh*(·) and *sigm*(·) are element wise non-linearities, with ⊙ representing element-wise multiplicaion. From a technical standpoint, attention based pooling allows different weights to be assigned to instances alleviating the need for explicit instance selection. Moreover, the final bag representation will be more informative. The synergistic combination of spatial and attention based pooling allows for improved training towards learning robust and interpretable features.

**Algorithm 1.**
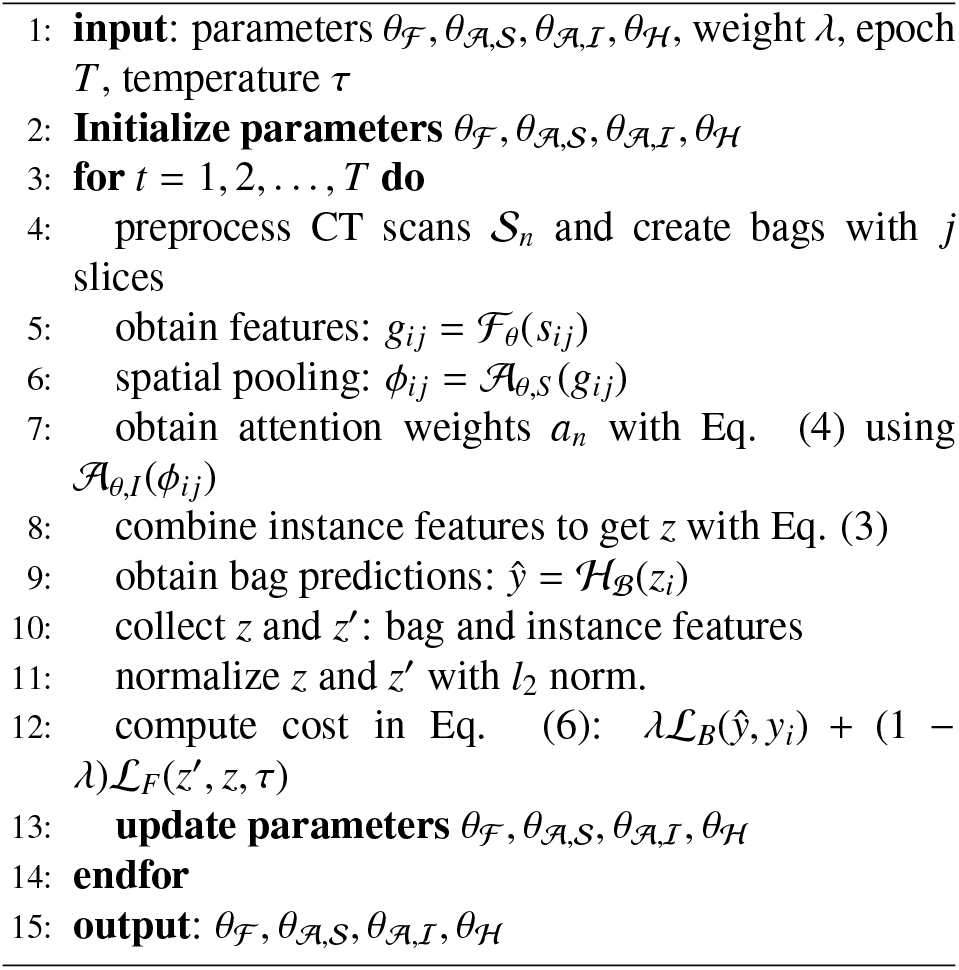
DA-CMIL Algorithm

#### 3.4. Contrastive MIL

Inspired by recently proposed self-supervised learning methodsChen et al. (2020a,b), we integrate unsupervised contrastive loss with the proposed MIL method for improved learning of instance level features. Formally, our model learns representations that maximize the agreement between instance features and aggregated bag features of the same patient via a contrastive lossChen et al. (2020a) in the latent space. Figure 2 shows the overall concept of the applied technique.

**Figure 2:**
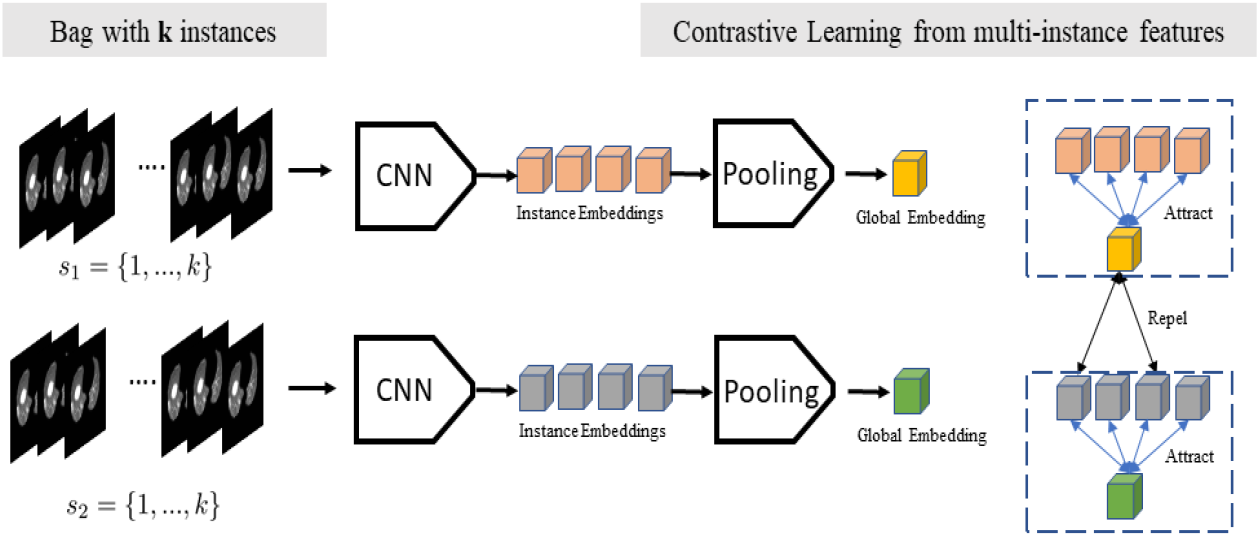
Illustration of contrastive learning applied to the MIL setting during model training.

According to the previously proposed self-supervised framework that uses contrastive loss, stochastic data augmentation is applied on 2D data samples to create two correlated views of the same example Chen et al. (2020b,a). Augmentations include random cropping, color distortions and random Gaussian bluring. Moreover, the contrastive loss is proposed to define contrastive predictive tasks on unlabeled samples, wherein positive and negative pairs are identified for given samples. To incorporate this idea, stochastic data augmentation is omitted in our study since contrastive loss is applied in the latent space. In addition, for any given patient CT scan; we infer that each slice can be considered as pseudo augmentation of the overall patient characteristics. Thus, we consider that the stochastic augmentation is implicitly applied (i.e. different views of the same patient).

Let *z*′ be the latent instance level feature of patient, and *z* the global patient-level aggregated feature obtained via the proposed modules. Then, following *l*_2_ normalization of *z*′ and *z* features, a contrastive loss can be defined as

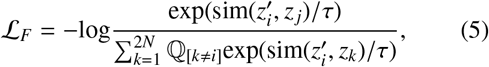

where 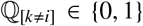 ∈ {0, 1} is an indicator function that evaluates to 1 iff *k* ≠ *i* and τ denotes a temperature parameter. *sim*(·, ·) is a similarity function i.e. cosine similarity. The loss is computed across all patient slice features and respective global features, herein considered as augmentations per mini-batch. The final loss function of the entire framework is defined as:

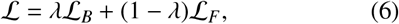

where λ is a parameter to weight the contribution of bag and constrastive losses, respectively. The detailed algorithm is presented in Algorithm 1.

### 4. Experiments

We evaluate the proposed method on a recently collected dataset and compare diagnostic performance against existing methods similar to ours. We present details on evaluation settings and any pre-processing applied.

#### 4.1. Datasets

In this study, we collected a chest CT dataset comprised of 173 samples at Yeungnam University Medical Center (YUMC), in Daegu, South Korea. The dataset includes 75 CT examples for patients with COVID-19, and 98 examples from patients with bacterial pneumonia collected between February and April, 2020. The study was approved by the Institutional Review Board (IRB) of Yeungnam University Hospital. COVID-19 patients were confirmed by RT-PCR assay of nasal and pharyngeal swab samples.

Further, we designed variants of YUMC CT dataset to fairly assess the performance of our method and others such as 3D based approaches. Namely, based on the original YUMC CT data using CT slices per patient, we processed a patch-based version of the dataset. In the MIL framework, 2D CT slice or patches can be used as instances, thus we evaluate our method on both cases. In addition, a 3D CT volume dataset is also processed for training/testing 3D based methods under fully-supervised settings.

For pre-processing, lung regions were segmented for all CT examples. To achieve this, we employed a ResNeSt Zhang et al. (2020a) model for segmentation training and inference. The model was trained on two public datasets i.e. non-small cell lung cancer (NSCLC) Aerts et al. (2014) and COVID-19 lung infection dataset Jun et al. (2020). Herein, a total of 50,756 lung slices were used for training and evaluated on 1,222 independent slices. Figure 3 shows examples of CT slices and patches employed.

**Figure 3:**
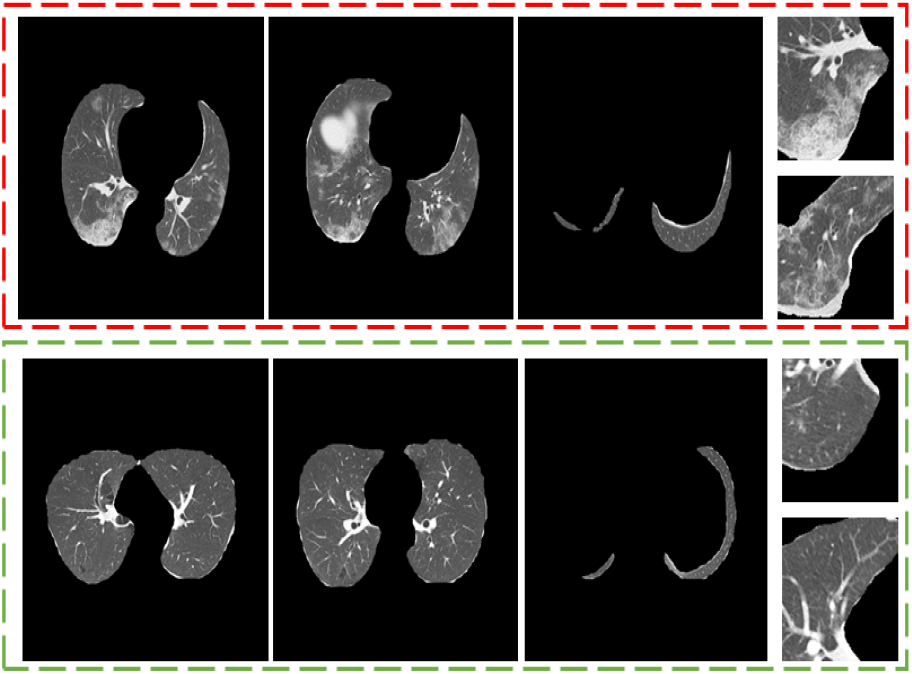
Pre-processed CT examples. (Red-section) COVID-19 CT slice and patch samples. (Green-section) bacterial pneumonia samples.

### 4.2. Experimental Settings

Accordingly, all the datasets were split into training, validation and testing by patient IDs with ratios 0.5, 0.1, and 0.4, respectively. The same split was used across all the dataset variants with all versions using only cropped lung regions. CT examples were 512×512, 128×128 and 256 × 256 × 256 in size for the slices, patches and 3D CT volume sets, respectively. Each CT slice was resized from 512×512 to 256×256 and patch slices were resized to 256 from 128. In particular, the slices set consisted of approximately 14,000 slices, whereas the patch version yielded 64,000 patches that mainly showed ≥ 30% of lung tissue. In the case of 3D CT volumes, all slices belonging to a patient were used to construct a volume with nearest neighbor sampling applied to obtain the desired input sizes.

The proposed model was implemented in Pytorch. A ResNet-34He et al. (2016) finetuned from imageNet pretrained weights was used as the feature extraction module 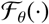, with a single fully connected (FC) layer employed as the bag classifier 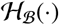. The dimension of the features was fixed to 512; this includes the feature maps obtained from 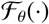 which had 512×8×8, with *C* = 512. Following spatial pooling, features were reshaped back to 512.

During training, data augmentation consisting of random transformations such as flipping were applied for both 2D and 3D based methods. All models were trained for 30 epochs except 3D based methods with an initial learning rate of 1_∊_ − 4 for 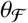, and 1_∊_ − 3 for the other modules with Adam optimization and a batch size of 8. On the other hand, 3D CNN based methods used a batch size of 4, learning rate of 1_∊_ − 4 and were trained for 60 epochs.

For the proposed method, a bag was constructed with *k* instances during training following step 4 of our algorithm, though for inference all available instances per patient were used to obtain the final prediction. We also evaluated the efficacy of our method based on varying *k* during training via ablation studies. For stable training, the learning rate was annealed by a factor of 0.1 at epochs 10, 15 and 25, respectively. We empirically set the loss hyper-parameters λ and τ to 0.5 and 1.0, respectively.

## 4.3. Comparison Methods

To evaluate the efficacy of the proposed method, we compared against recent MIL based methods i.e. Deep-AttentionMIL Ilse et al. (2018), ClassicMIL Campanella et al. (2019) and JointMILChikontwe et al. (2020). Also, recent 3D based methods DeCovNetWang et al. (2020c) and Zhang3DCNNZhang et al. (2020b) were included for comparison. For a fair evaluation, the same backbone feature extractor is used in all methods except for the 3D methods as we used the publicly available implementations.

In particular, ClassicMIL follows the traditional assumption of the MIL setting and focuses on instance level learning wherein only the top instance per bag is considered for the final patient level prediction. DeepAttentionMIL uses attention-based pooling for bag level learning. In constrast, JointMIL combines both instance and bag level learning with bag feature clustering during training. Lastly, DeCoVNet and Zhang3DCNN both use all available CT slices in a constructed volume under the fully supervised setting. The later methods serve as an upper-bound over the weakly supervised methods evaluated in this study.

## 5. Results

We present both quantitative and qualitative results of the proposed methods. Also, ablation studies on the effect of bag size and the weighting parameter λ are presented.

### 5.1. Quantitative Results

Diagnostic performance was evaluated on YUMC CT slice, patch and CT volume based datasets using accuracy, area under the curve (AUC), f1-score, specificity and sensitivity, respectively. Tables 1 and 2 show the performance of the evaluated methods on the datasets.

**Table 1:** Evaluation of the proposed methods on YUMC dataset including results of using D*A*-CMIL with/without contrastive loss 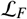.

| Method | Accuracy | AUC | F1 | Specificity | Sensitivity |
| --- | --- | --- | --- | --- | --- |
| DeCoVNet Wang et al. (2020c) | 0.831 | 0.825 | 0.8 | 0.875 | 0.774 |
| MIL Campanella et al. (2019) | 0.803 | 0.796 | 0.767 | 0.85 | 0.742 |
| DeepAttentionMIL Ilse et al. (2018) | 0.859 | 0.875 | 0.861 | 0.75 | 1 |
| JointMIL Chikontwe et al. (2020) | 0.901 | 0.909 | 0.896 | 0.85 | 0.968 |
| Zhang3DCNN Zhang et al. (2020b) | 0.93 | 0.938 | 0.925 | 0.875 | 1 |
| DA-CMIL (w/o $\mathcal{L}_F$) | 0.93 | 0.934 | 0.923 | 0.9 | 0.968 |
| DA-CMIL (w/ $\mathcal{L}_F$) | **0.986** | **0.984** | **0.984** | **0.975** | **1** |

**Table 2:** Evaluation of the proposed methods on YUMC patch dataset.

| Method | Accuracy | AUC | F1 | Specificity | Sensitivity |
| --- | --- | --- | --- | --- | --- |
| MIL Campanella et al. (2019) | 0.845 | 0.852 | 0.836 | 0.8 | 0.903 |
| DeepAttentionMIL Ilse et al. (2018) | 0.845 | 0.859 | 0.845 | 0.75 | 0.968 |
| JointMIL Chikontwe et al. (2020) | 0.845 | 0.837 | 0.814 | 0.9 | 0.774 |
| DA-CMIL (w/o $\mathcal{L}_F$) | 0.93 | 0.934 | 0.923 | 0.9 | **0.968** |
| DA-CMIL (w/ $\mathcal{L}_F$) | **0.958** | **0.955** | **0.951** | **0.975** | 0.935 |

In Table 1, D*A*-CMIL with contrastive loss 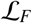 achieves the best overall performance of 98.6% accuracy and an AUC of 98.4%. Notably, even when 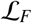 was not applied during training, our method still reports 93%(+2.9) and 93.4%(+2.5) in terms of accuracy and AUC over the best weakly supervised method JointMIL. MIL reports the lowest performance among all methods, which is expected since it only considers the top instance among multiple 2D slices in bag for inference. Interestingly, our method outperformed both Zhang3DCNN and DeCoVNet which are fully supervised methods even without 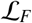 used in the training stage. Though both D*A*-CMIL and DeepAttentionMIL employ attention based pooling, the proposed method shows improved performance via dual-attention pooling, validating the architectural design.

To further validate the proposed method, we applied D*A*-CMIL to randomly cropped patches of the CT samples. As shown in Table 2, performance was consistently better than the compared methods. All weakly supervised method showed similar accuracy with considerable margins observed for sensitivity. DeepAttentionMIL reported the best sensitivity at 96.8% with accuracy consistent with other methods. However, D*A*-CMIL showed an improvement of +11.3% in accuracy over the best compared method with an equally larger margin without 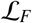 employed. The effect of using attention and contrastive loss was more pronounced in the case of patches as not applying 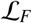 showed reduced performance (−2.8%).

Figure 4 shows the receiver operating characteristic(ROC) curves of the compared methods on different datasets. Overall, the proposed method shows a high TPR and lower FPR across all settings. This is further evidenced in the summaries of the confusion matrices of the comparison methods as presented in Figures 5 and 6. This indicates D*A*-CMIL can be viable option for accurate and robust screening of COVID-19.

**Figure 4:**
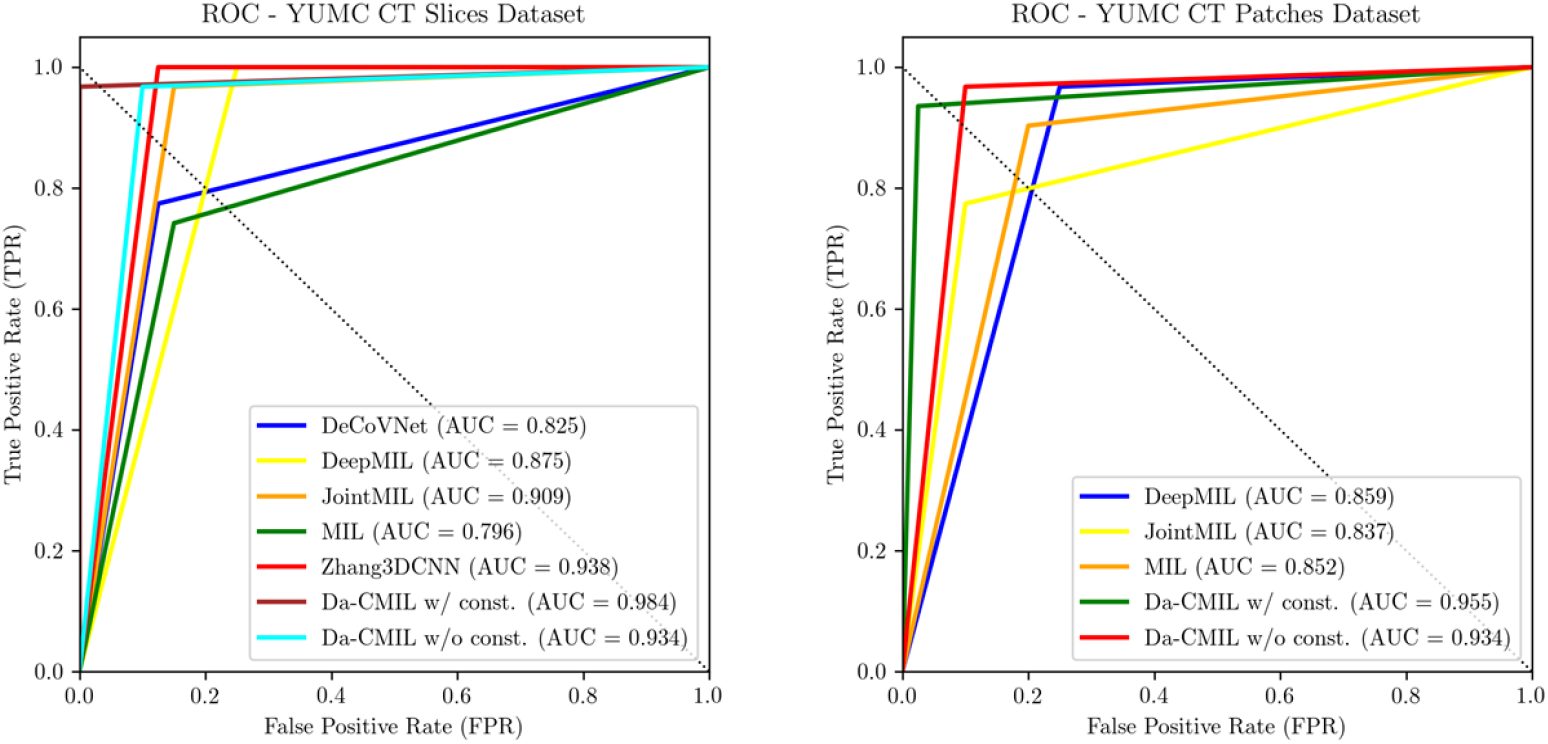
The Receiver operating characteristic(ROC) curves of compared methods on the YUMC CT slices and patch datasets.

**Figure 5:**
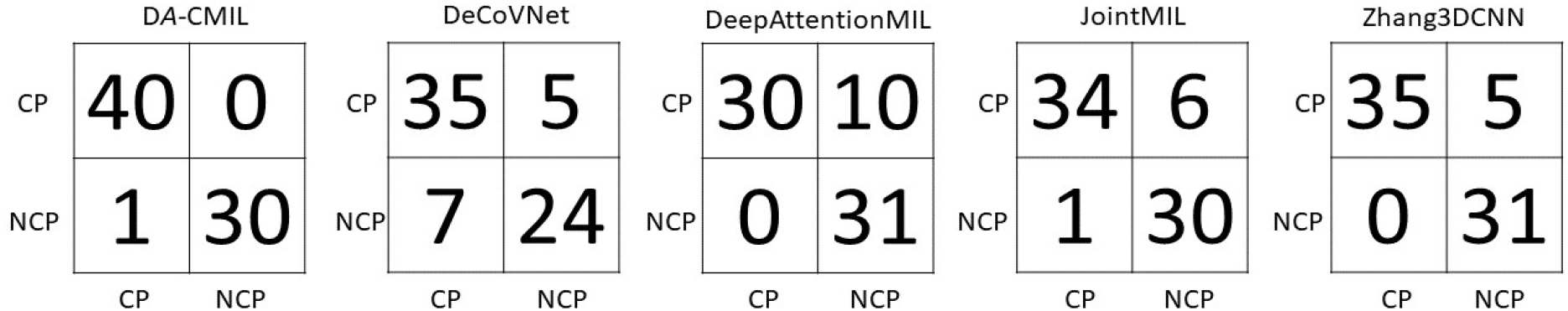
Confusion matrices of compared methods on YUMC CT Slices dataset. CP represent Pneumonia and NCP implies COVID-19, respectively.

**Figure 6:**
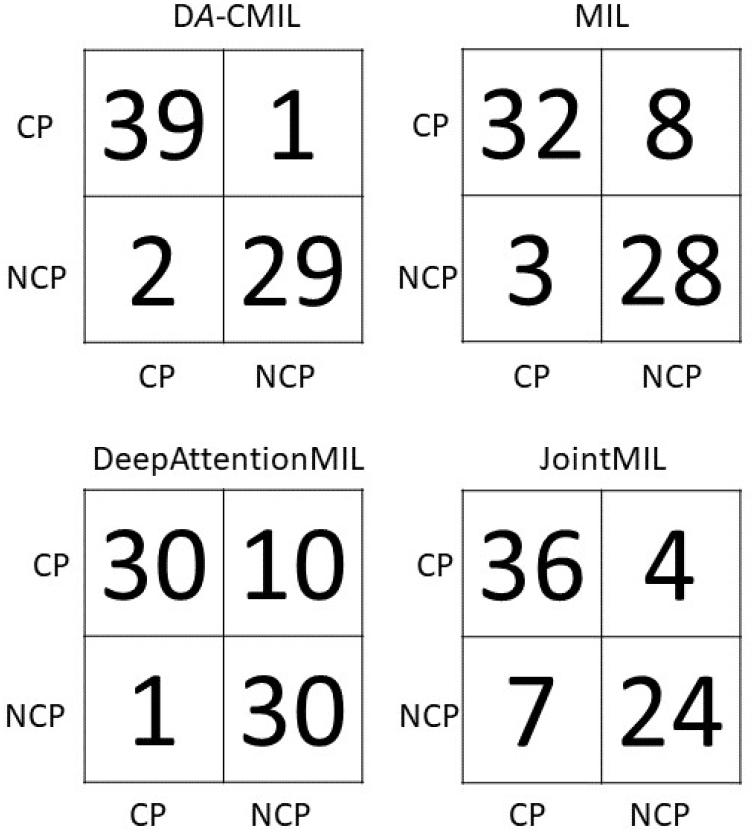
Confusion matrices of compared methods on YUMC CT patch dataset.

### 5.2. Effect of the bag size

To assess the effect of bag size during training on the proposed method, we performed an ablation study where the bag was constructed by varying *k* i.e. each bag consisted of *k* max instances (slices/patches). As shown in Table 3 and Figure 7, as the bag size increases D*A*-CMIL performance improves. The best result was achieved when *k* = 32 with a considerable margin across all metrics. We limited evaluation to *k* = 32 due to computational limitations. Moreover, it worth noting that contrastive methods benefit from large batch sizes; as evidenced from the reported results, our findings are consistent with existing observations based on self-supervised methods applied to general vision datasets. However, as *k* is increased the relative batch size based on bags should be reduced to compensate for training time and memory requirements. In general, results show that our method is not limited/affected by the number of instances available per CT scan and can benefit from using more instances for training, though during the inference stage all instances are used.

**Table 3:** Evaluation of varying bag sizes with the proposed method on YUMC CT slices dataset.

| Method | Accuracy | AUC | F1 | Specificity | Sensitivity |
| --- | --- | --- | --- | --- | --- |
| DA-CMIL (w/ $k = 8$) | 0.93 | 0.934 | 0.923 | 0.9 | 0.968 |
| DA-CMIL (w/ $k = 16$) | 0.944 | 0.939 | 0.933 | 0.975 | 0.903 |
| DA-CMIL (w/ $k = 24$) | 0.944 | 0.943 | 0.935 | 0.95 | 0.935 |
| DA-CMIL (w/ $k = 32$) | **0.986** | **0.988** | **0.984** | **0.975** | **1** |

**Figure 7:**
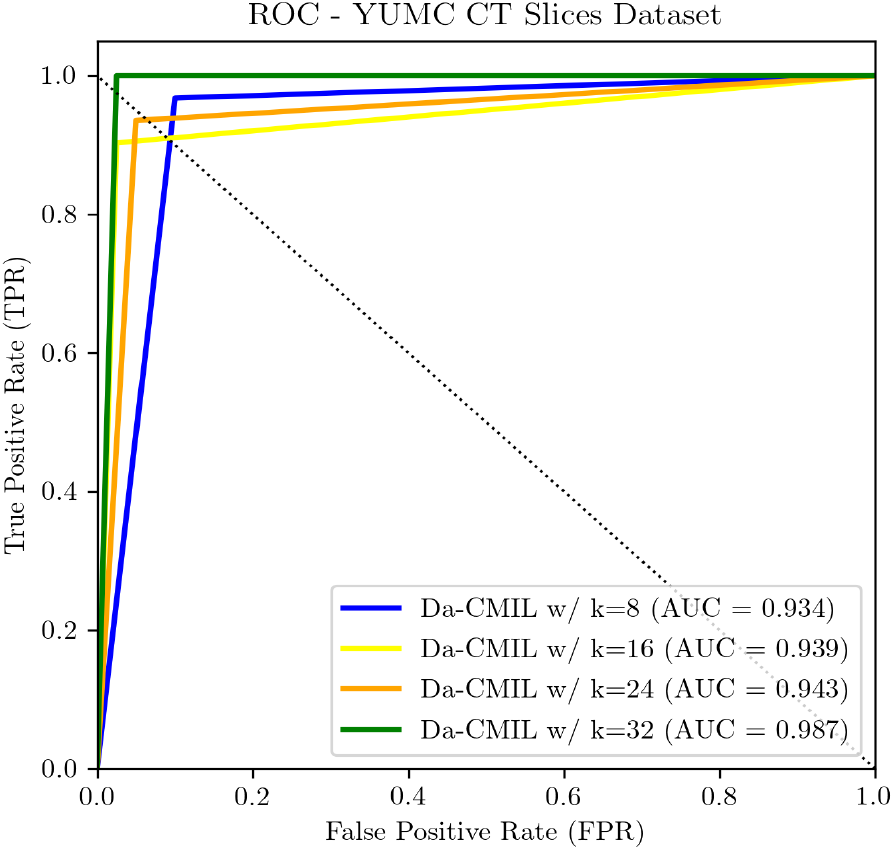
ROC curves of DA-CMIL on YUMC CT Slide dataset when *k* is varied during training.

### 5.3 Effect of the weight parameter λ

D*A*-CMIL uses contrastive feature learning of multiple instances with a weighting parameter λ to balance the effect of the losses. When λ = 1.0, 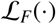 has no effect on learning and showed a lower performance of 93% compared to not using 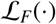 i.e. when λ < 1.0. Though similar performance was noted across different values of λ, the most significant is when contrastive loss was not applied entirely as presented in Table 4.

**Table 4:**
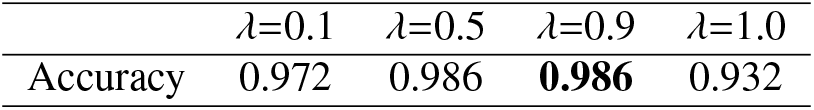
Evaluation on varying λ in the cost function 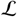 on the YUMC Dataset.

### 5.4. Qualitative Results

In Figure 8, qualitative results are presented based on spatial attention maps and attention scores, respectively.

**Figure 8:**
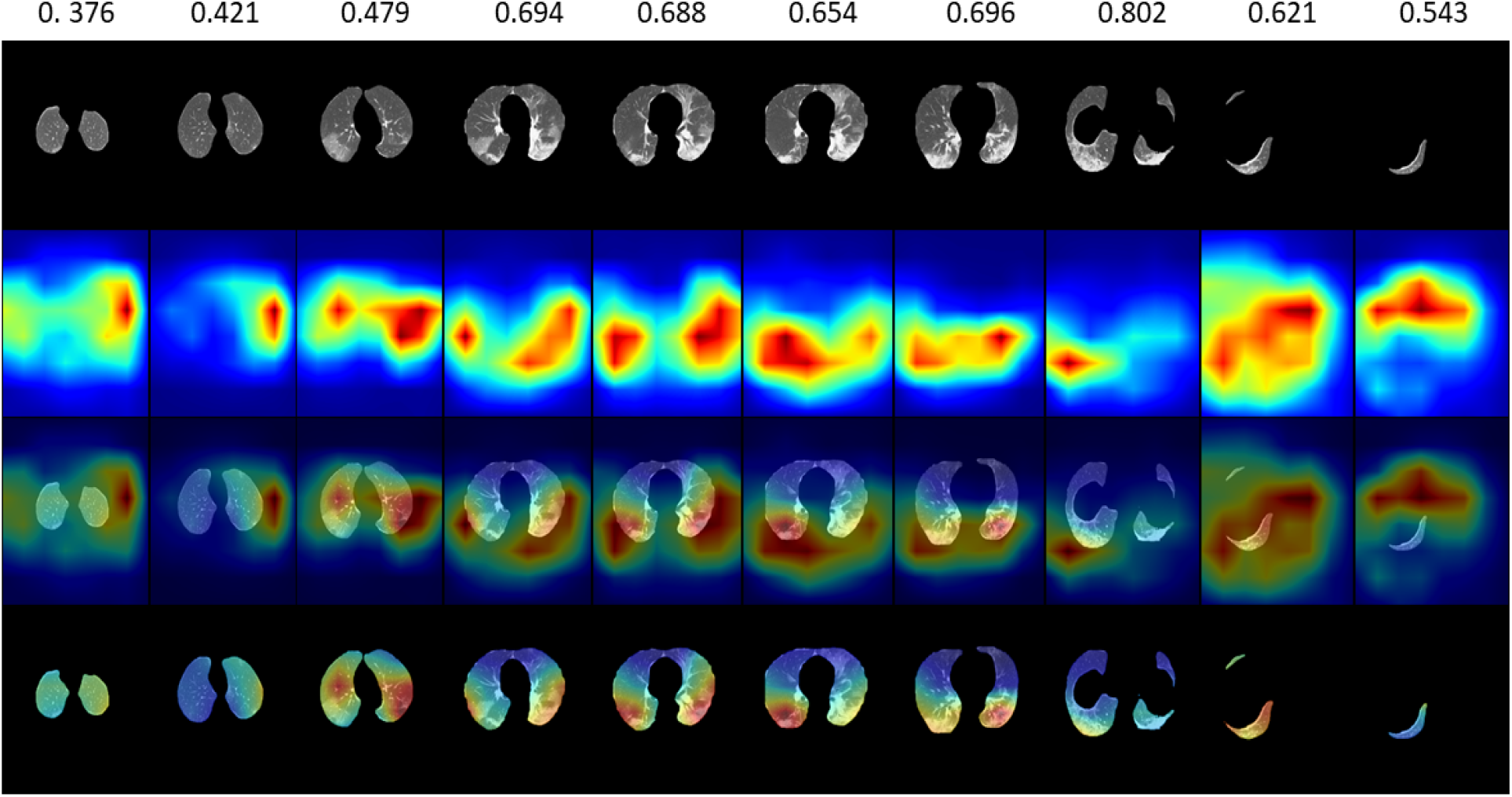
Qualitative examples of D*A*-CMIL spatial attention maps with attention scores on CT samples from a single patient with COVID-19. The top row shows the attention value of each slice with the spatial maps normalized to focus on the lung regions only.

This demonstrates that D*A*-CMIL is able to find key slices related to infected areas with coarse maps. Interestingly, low attention scores were observed for slices such as noisy slices/artifacts with no infected areas further indicating the utility of our method. Moreover, attention maps focus on key areas such as ground-glass opacities and consolidations, both consistent with clinical findings. This can be highly beneficial in clinical evaluation since our method avoids post-hoc analysis based on class activation maps (CAM).

## 6 Discussion

Though RT-PCR is the gold standard for COVID-19 diagnosis, it is still hindered by lengthy test times, as it can take days to obtain the results. Accordingly, CT has been considered as a reasonable alternative for current testing methods as it can produce results within minutes. We showed a novel approach to the application deep CNNs for COVID-19 diagnosis under weak supervision with clinical implications. It is important to have a fully automated and interpretable method in actual settings for rapid evaluation. Moreover, given the subtleties that exist between COVID-19 and other pneumonia in terms of imaging characteristics that field experts find hard to differentiate, accurate diagnosis is highly relevant.

Our method was evaluated on recently curated dataset wherein only patient diagnostic labels are available without lesion infected regions of interest as is common in existing methods. To further validate our approach, we qualitatively showed the regions that are focused on by our model via coarse attention maps alongside attention scores. Our method achieved an AUC of 98.4%, accuracy of 98.6% and a true positive rate (TPR) of 96.8%. In addition, attention maps obtained highlight key infection areas in the majority of samples with attention scores corresponding to key slices.

We also empirically showed the benefit of using an unsupervised contrastive loss to complement the supervised learning of patient labels and may serve as a base for more complex methods. Moreover, the proposed method surpassed 3D based methods by large margins. We infer this may be due to the limited size of the dataset employed as most recent methods applied to 3D CT volumes report using large cohorts in literature. In addition, since DeCoVNet was trained from scratch and has an custom deep architecture, performance was subpar. Though ZhangCNN’s performance was considerably better than the later, it still did not achieve comparable performance even when the model was trained for more epochs. It is also worth noting that models trained with extensive augmentation did not achieve any considerable improvements across the evaluation metrics, since COVID-19 and bacterial pneumonia present similar characteristics.

There exist a few limitations with regard to the proposed method. Though attention maps could show interpretability and explainability for COVID-19 diagnosis, there exist some failure cases where the attention map do not correctly indicate an infected region as shown inFigure 5. Second, we found that extensive data augmentation such as color jittering lead to reduced performance and was largely negligible compared to the benefit of using a contrastive loss which showed consistent improvements across all evaluation settings. This motivates us to consider using more complex attention modes for better diagnostic interpretability as well as explore unsupervised pre-training using the proposed method both in 2D or 3D as future directions.

## 7. Conclusion

In this study, we developed a 2D CNN framework with dual attention modules and contrastive feature learning under the multiple instance learning (MIL) framework to distinguish COVID-19 and a bacterial sub-type of pneumonia in chest CTs. We verified performance on both CT patch and slice based versions of the datasets and report results comparable to state-of-the-art methods. In addition, ablation experiments show the benefit of using large bag sizes during training and the effect of weighting losses correctly for stable learning. Through this study, we hope to add valuable contribution to the current literature on weakly supervised methods for COVID-19 screening.

### CRediT authorship contribution statement

**Philip Chikontwe**: Conceptualization, Formal analysis, Investigation, Methodology, Validation, Visualization, Software, Writing - original draft, Writing - review & editing. **Miguel Luna**: Investigation, Methodology, Writing - review & editing. **Myeong Kyun Kang**: Investigation and Methodology. **Kyung Soo Hong**: Resources & Data curation. **June Hong Ahn**: Resources, Data curation, Supervision, Writing - review & editing. **Sang Hyun Park**: Funding acquisition, Methodology, Project administration, Supervision, Writing - review & editing.

## Data Availability

Data will be available following publication.

## Acknowledgments

This work was supported by the National Research Foundation of Korea (NRF) grant funded by the Korean Government (MSIT)(No.2019R1C1C1008727).

